# Paired nasopharyngeal and deep lung testing for SARS-CoV2 reveals a viral gradient in critically ill patients: a multi-centre study

**DOI:** 10.1101/2020.07.19.20156869

**Authors:** Islam Hamed, Nesreen Shaban, Marwan Nassar, Dilek Cayir, Sam Love, Martin D Curran, Stephen Webb, Huina Yang, Katherine Watson, Anthony Rostron, Vilas Navapurkar, Razeen Mahroof, Andrew Conway Morris

## Abstract

Samples for diagnostic tests for SARS-CoV-2 can be obtained from the upper (nasopharyngeal/oropharyngeal swabs) or lower respiratory tract (sputum or tracheal aspirate or broncho-alveolar lavage - BAL). Data from different testing sites indicates different rates of positivity. Reverse-transcriptase polymerase chain reaction (RT-PCR) allows for semi-quantitative estimates of viral load as time to crossing threshold (Ct) is inversely related to viral load.

**Objectives:** The objective of our study was to evaluate SARS-CoV2 RNA loads between paired nasopharyngeal (NP) and deep lung (endotracheal aspirate or BAL) samples from critically ill patients.

**Methods:** SARS-CoV-2 RT-PCR results were retrospectively reviewed for 51 critically ill patients from 5 intensive care units in 3 hospitals ; Addenbrookes Hospital Cambridge (3 units), Royal Papworth Cambridge (1 unit), and Royal Sunderland Hospital (1 unit). At the times when paired NP and deep lung samples were obtained, one patient had been on oxygen only, 6 patients on non-invasive ventilation, 18 patients on ECMO, and 26 patients mechanically ventilated.

**Results:** Results collected showed significant gradient between NP and deep lung viral loads. Median Ct value was 29 for NP samples and 24 for deep lung samples. Of 51 paired samples, 16 were negative (below limit of detection) on NP swabs but positive (above limit of detection) on deep lung sample, whilst 2 were negative on deep sample but positive on NP (both patients were on ECMO).

**Conclusions:** It has been suggested that whilst SARS-CoV1 tends to replicate in the lower respiratory tract, SARS-CoV2 replicates more vigorously in the upper respiratory tract. These data challenge that assumption. These data suggest that viral migration to, and proliferation in, the lower respiratory tract may be a key factor in the progression to critical illness and the development of severe acute respiratory syndrome (SARS). Factors which promote this migration should be examined for association with severe COVID-19. From a practical point of view, patients with suspected severe COVID-19 should have virological samples obtained from the lower respiratory tract where-ever possible, as upper respiratory samples have a significant negative rate.

---

Since the start of the SARS-CoV2 pandemic, approximately 13000 patients have been admitted to critical care in the United Kingdom, the majority have required advanced respiratory support^1^ Samples for SARS-CoV-2 detection can be obtained from the upper (nasopharyngeal/oropharyngeal swabs) or lower respiratory tract (sputum/endotracheal aspirate/broncho-alveolar lavage (BAL))^2^. Viral RNA is detected using reverse transcriptase polymerase chain reaction (RT-PCR). The Cycle threshold (Ct) has a simple negative linear correlation with the logarithm of the number of gene copies in the original sample and thus can be used to provide a semi-quantitative estimate of the viral RNA in a specimen^3^.

It has been suggested that SARS-CoV-2 is predominantly shed from upper respiratory tract, distinguishing it from SARS-CoV-1, where replication occurs mainly in the lower respiratory tract.^4-6^ A recent multi-site viral detection study^5^ indicated higher nasopharyngeal (NP) viral loads in some patients early in the course of disease, although they generally detected viral RNA in sputum for longer. However, this study^5^ was conducted on patients with mild disease, and it is unclear whether the results pertain to critically ill patients.

Our objective was to evaluate SARS-CoV2 RNA loads between paired NP and deep lung (endotracheal aspirate or BAL) samples from critically ill patients.

## Methods

Patients admitted to five intensive care units (ICU) in three UK hospitals with PCR-confirmed COVID19 were identified retrospectively. The sites were: Addenbrookes Hospital Cambridge, a tertiary centre with 3 units (a general ICU, Neuro-trauma unit and dedicated COVID unit), Royal Papworth Hospital Cambridge (tertiary respiratory failure and extra-corporeal membrane oxygenation (ECMO) unit), and Sunderland Royal Hospital (general unit) a large district general hospital. Patients with paired NP and deep lung samples were identified, with ‘paired’ defined as samples taken within 24 hours of each other. At Addenbrooke’s and Royal Papworth, samples were analysed in a common microbiology laboratory, using in-house RT-PCR following extraction using the Easy Mag platform (Biomeriuex, Basingstoke, UK). Samples from Royal Sunderland were analysed using the Cobas 6800 (Roche, Welwyn, UK). Clinical data was obtained from case note review. As a retrospective service evaluation of anonymised routinely collected data, the requirement for consent was waived.

## Results

51 patients with paired samples were identified. Median age of patients was 49 years (range 24-74), with 74% males. The median duration between onset of symptoms and when paired samples were obtained, was 10 days (range 1-22). When paired samples were obtained, one patient was receiving oxygen via a simple facemask, 6 patients were receiving non-invasive ventilation or mask CPAP, 26 patients mechanically ventilated and 18 were on ECMO.

There was a significant gradient between NP and deep lung viral loads (Figure 1) with median Ct value of 29 for NP and 24 for deep lung samples. Of 51 paired samples, 16 were negative on NP swabs but positive on deep sample, whilst 2 were negative on deep sample but positive on NP (both patients were on ECMO). Of the subgroups by ventilatory support, the 26 mechanically ventilated patients demonstrated the largest gradient, median Ct value was 28 for NP samples and 19 for deep lung samples, these were also the patients with the shortest interval between symptom onset and testing (median 5 days range 1-11).

**Figure 1:**
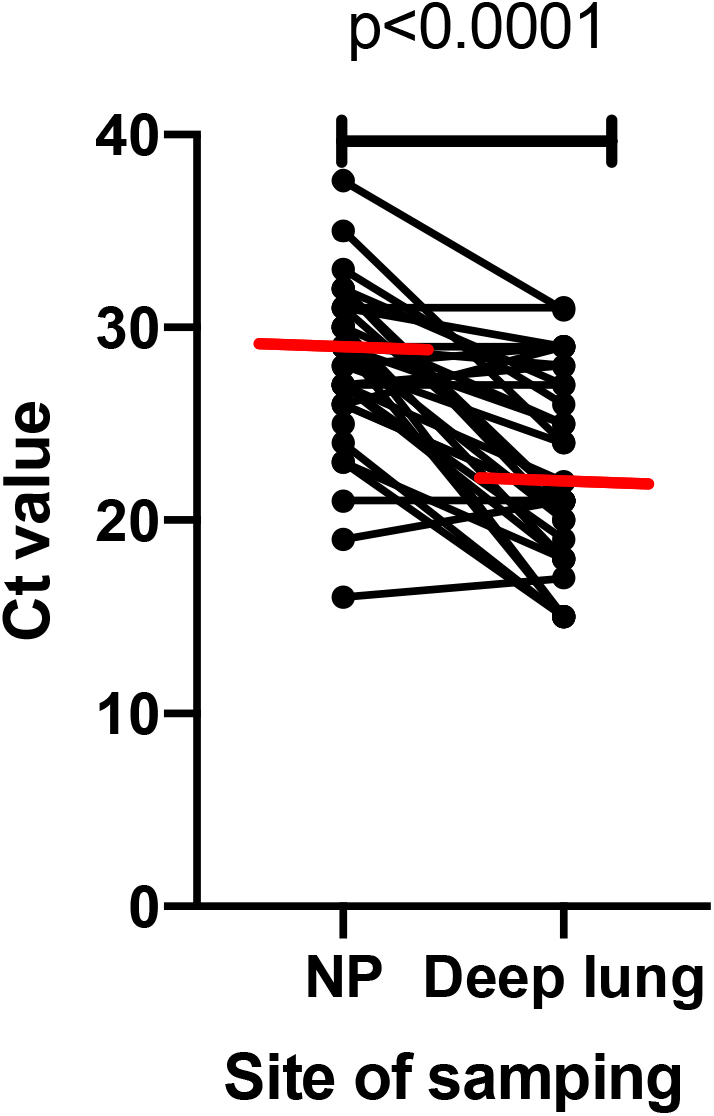
Comparison of viral load of paired samples from nasopharynx (NP) and deep lung (endotracheal aspirate or BAL), excluding samples where NP (16) or deep lung (2) were below the limit of detection. Red line indicates median value. P value by Wilcoxon matched pairs test

## Discussion

In our case series of critically ill patients with COVID19, NP swabs were relatively insensitive for detection of SARS-CoV2: 69% of NP samples detected viral RNA compared to 96% of deep respiratory samples. There was also a clear viral gradient with a median 5 cycle (9 cycles for mechanically ventilated) lower Ct value in the lungs. To the best of our knowledge, this is the first report of paired respiratory samples from critically ill patients with COVID19.

In SARS, arising from SARS-CoV1, there was a substantial rate of false negative nasopharyngeal swabs^6^ leading to the suggestion that SARS-CoV1, unlike SARS-CoV2, had a predilection for lower airways^4^. Our work challenges this assumption, demonstrating significantly higher viral loads in the lower respiratory tract amongst critically ill patients. It is possible that the higher viral load in the lungs may contribute to the harmful inflammatory response that constitutes the pathology of SARS, although recent post-mortem reports indicate that pulmonary clearance of the virus does not lead directly to resolution of inflammation^7^. Wherever possible, our data suggest that in patients with suspected severe COVID19, deep respiratory samples should be taken to improve diagnostic yield. This will become increasingly important as the Northern Hemisphere enters the winter and the prevalence of other viral pneumonias increases. Although there is understandable concern about the risk of deep lung sampling such as BAL or endotracheal aspirate generating aerosol and increasing risk of healthcare worker infection, our experience during the pandemic^8,9^ is that such procedures can be performed safely. Use of enhanced aerosol protecting personal protective equipment, respiratory isolation and closed/semi-closed respiratory circuits combine to increase the safety of this process.

This study reports from five units using two different molecular tests, increasing the generalisability of our findings. We acknowledge that the retrospective nature of the data collection may introduce a source of bias against NP testing, which is often used as the first line test, and that deep respiratory sampling may have only been undertaken if RNA was not detected from NP swabs. However, during the first wave of the pandemic, the turn-around time for PCR was generally greater than 24 Hours, and it is unlikely that negative NP swabs will have influenced the decision to obtain paired samples within the same 24 hour period. Although all NP swabs were taken by nurses appropriately trained in viral sample acquisition, we cannot be certain that the patient’s nasopharynx was correctly sampled in all cases, however it does reflect the real-world experience of virological sampling.

In conclusion, we have found that critically ill patients with COVID19 demonstrate a significant viral gradient from the upper to the lower respiratory tract, which may have diagnostic and pathophysiological importance. We conclude that, wherever possible, lower respiratory tract samples should be obtained for the detection of SARS-CoV2.

## Data Availability

data refers to individual patients and is not available for general use.

